# Periodic epidemic outbursts explained by local saturation of clusters

**DOI:** 10.1101/2022.08.31.22279430

**Authors:** Louis Gostiaux, Wouter J. T. Bos, Jean-Pierre Bertoglio

## Abstract

Adding the notion of spatial locality to the susceptible-infected-recovered (or SIR) model, allows to capture local saturation of an epidemic. The resulting minimum model of an epidemic, consisting of five ordinary differential equations with constant model coefficients, reproduces slowly decaying periodic outbursts, as observed in the COVID-19 or Spanish flu epidemic. It is shown that if immunity decays, even slowly, the model yields a fully periodic dynamics.

---

Mathematical models of epidemics remain indispensable in the fight against diseases such as the COVID-19 crisis. These models help to take political decisions such as mobility restrictions. An example of such a model is the compartmental SIR^1^ model, which is the paradigm-model to illustrate the elementary dynamics of epidemic spreading in a well-mixed community.

The SIR dynamics describe the exponential increase of infected individuals upto a level where the epidemic saturates through herd-immunity. After this saturation, the number of infected individuals declines exponentially towards zero. In most epidemics, this global immunity is not attained after the first peak, or wave of infections. Indeed, another feature of epidemics is quasi-cyclic behavior with several waves of infection spaced by a period of several months. This behavior is for instance observed during the COVID-19 pandemia^2^, the 2014 Ebola outbreak in Guinea^3^ as well as during the influenza epidemic in the early 20th century^4^.

The main contribution of the present work is the insight that these two features (rapid saturation and quasi-cyclic behavior) are caused by the same underlying physical phenomenon. We propose a novel modeling of the global effect of local epidemic saturation. To this end, we introduce the concept of local herd immunity, which means that, in the population directly in contact with the infected individuals (or clusters), a sufficient amount of individuals is immune, so that spreading slows down or even stops^5^. This does in general not imply that (global) herd-immunity is attained by the total population. To understand how exactly this local herd immunity leads to epidemic waves, we derive and analyse a novel variant of the SIR model, introducing the minimum amount of additional complexity required to obtain successive epidemic waves.

The SIR model is too simple to describe the dynamics of a realistic epidemic, in particular since no distinction is made between susceptible individuals which are in contact, or on the contrary, far away from contagious individuals. This well-mixedness assumption can be leveraged in various ways. One possible way is to radically change the approach and consider agent-based descriptions^6–8^. If one wants to keep a compartmental approach, diffusion can be added to the system^9,10^, memory-effects^11^, or one can consider SIR-type models on lattices or using small-world networks^12–14^. We recently proposed a minimal refinement to introduce the notion of space-dependence in the compartmental approach, dividing the community into only two groups: individuals close to, and those far away from the infected^5^. It is this approach, which succesfully introduced the effect of local herd-immunity on the first wave of the epidemics, that we will improve in order to model successive outbursts.

In the domain of mathematical modeling of epidemics, various ways are explored to reproduce quasi-cyclic behavior^15^. Examples are to adapt predator-prey descriptions^16,17^ or to introduce time-dependent model constants^18–20^. This latter approach needs to arbitrarily set the period and the beginning of the pandemic. However, the emergence of pandemics is in general not correlated to typical seasons, and can occur at any time^21^. What is new here is that we do not explicitly try to model the quasi-cyclic behavior; instead we demonstrate that it is a direct consequence of the spatial dynamics of an epidemic. The present investigation suggests that saturation is attained well before global collective immunity is reached, due to local collective immunity in clusters, and that after a phase of decline, the epidemic revives when local outbreaks have decayed sufficiently to become mathematically equivalent to new small-size clusters.

## a. SBIGR, the global dynamics of an epidemic in a non-fully-mixed community

In the following we introduce the SBIGR approach, which is a compartmental model. The acronym comes from the names of the five compartments that we will introduce, as usual in compartmental modeling. In addition to the classical *S, I, R* susceptible, infected, recovered (or removed) compartments, there are two new compartments *B, G*, which introduce the notion of spatial locality in the model. We now explain this further.

The total population consists of *N* individuals. Within well-defined clusters the local dynamics is given by a SIR like approach. We define a sub-ensemble *N*\* of individuals within lo-cal clusters around the infected. These individuals are either (local) susceptible *B*, infected *I* or (local) recovered *G*. For the individuals far away from the infected, we have in our desciption two types: susceptibles *S* and recovered *R*. Both the ensembles *S* and *B* indicate therefore susceptibles, far away from, or close to the infected, respectively. Similarly *R* and *G* both indicate recovered individuals. It is important to understand that the sub-ensemble *N*\*, in the following named the blob, represents multiple independent clusters with little or no connectivity, that will share similar constant physiological and social parameters, leading to similar dynamical equations. Both *N* and *N*\* are thus macroscopic quantities. The 5 types of individuals are represented as compartments in Fig. 1.

**FIG. 1.**
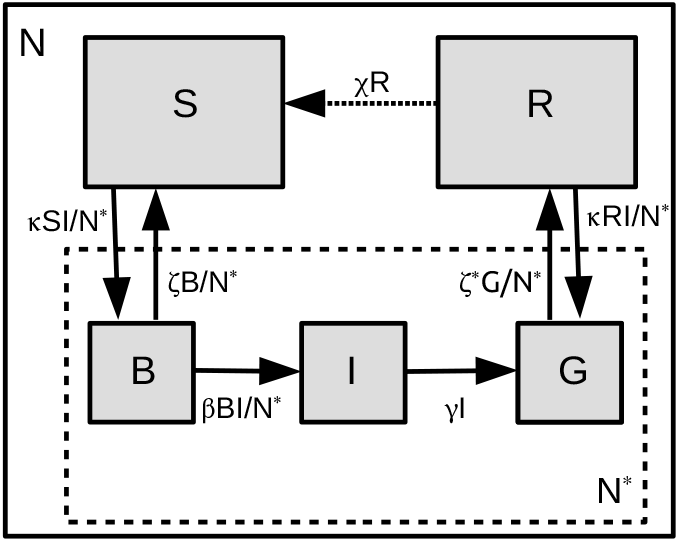
Representation of the SBIGR model. The susceptible part of the population is subdivided into individuals *B* inside the blob, and *S*, outside the blob. The recovered are also subdivided into individuals *G* inside and individuals *R* outside the blob. The blob population is constituted by *N*\* = *B* + *I* + *G*. The dashed arrow from the *R* to the *S* compartment reflects the temporal decrease of immunity.

Within the blob, corresponding to the ensemble of clusters, we have a SIR type of dynamics,

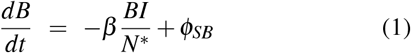

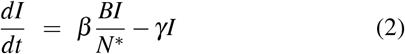

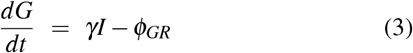

with *N*\* = *B* + *I* + *G*. For a fixed total population size, we have then

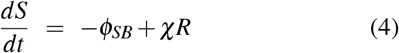

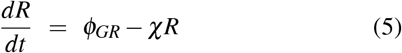

where we have added an exchange term *χR* from the *R* to the *S* compartment. This term represents the decrease of immunity after infection, that we will consider null (*χ* = 0) for the moment. Without the fluxes *ϕ*_*SB*_ and *ϕ*_*GR*_ we have a local SIR system, with *B, G* taking the place of *S, R*, respectively. Schematically, the compartments are shown in Fig. 1, where the fluxes *ϕ*_*SB*_ and *ϕ*_*GR*_ are indicated by arrows in both directions. The novelty of the SBIGR model is to estimate these fluxes with a physical model for the spatial evolution of the clusters, introducing a diffusion approach for the blob evolution^22^.

In Fig. 2 we propose a visual representation of the model. We insist here that this is not a spatial representation of clusters and moving individuals, but a schematic where the ensemble of clusters is regrouped in one single blob. We can then consider that the size of the blob evolves but that the individuals remain fixed in space. The collective effect of the movement of individuals within the blob, resulting in contamination by contact, is embodied by the value of *β*. The effect of interactions between individuals which are inside and outside a cluster determines the values of *κ* and *ζ*^5^.

**FIG. 2.**
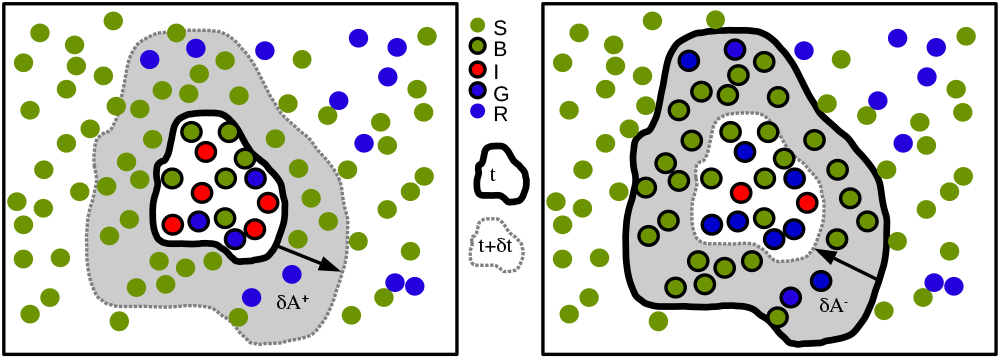
Graphical representation of the evolution of an (a) accelerating, or (b) decelerating epidemic. The “blob” around the infected *I* represents all people that are in close contact with contagious individuals. The blob expands when the local concentration of *I* is large. The blob will shrink if little infected are present. *S* and *B* are susceptibles, *G, R* are recovered. Change in surface, *δ A* during a time-interval *δt* is indicated as a shaded area.

This approach allows a geometrical interpretation of the present compartmental model without introducing an agent-based description. In this framework, we do not resolve the movement of individuals explicitly, but describe the evolution of the fictive surface of the blob. We call this surface *A*\* and the total surface containing all individuals *N* is denoted *A*. We have then that *N*\**/N* = *A*\**/A*.

On the left of Fig. 2 we illustrate the blob expansion, governed by the proximity of infected people (inside the blob, close to the border) with susceptibles (outside the blob). In a time-interval *δt*, it is observed that the number of susceptible individuals, which were outside the blob but who will become part of it, is given by

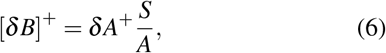

where *S/A* is the average concentration of susceptibles outside the blob. In the schematic, *δA*^+^ corresponds to the increase of surface of the blob. This increase of surface should be, at least dimensionally, proportional to 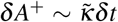, where 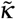 is a diffusion coefficient. The unknown at this point is how 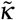 depends on the other parameters of the problem. What we do know is that the blob will expand when the concentration of infected individuals is large enough. We should therefore have a dependency on the local proportion of infected in the blob, *I/N*\*, and we have therefore in the simplest form 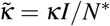 and thereby

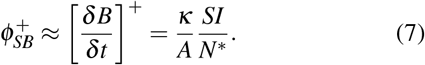

Let us now consider the case where the blob shrinks. Similarly, the amount of individuals that were in the blob, but will find themselves outside it, are given by the decrease of surface, multiplied by the local concentration of the *B*. We have therefore, if the blob shrinks, that

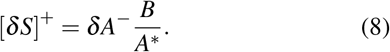

If we now model the shrinking by a negative diffusion process, independent of the presence or not of infected people, and we introduce *ζ* the diffusion coefficient associated with this process, we have

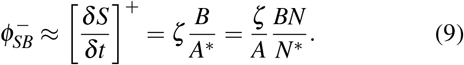

Thereby we have modeled both the positive and the negative contributions to the flux between *S* and *B*,

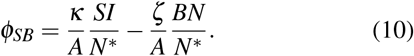

Exactly the same derivation, considering now the *G, R* individuals, leads to the expression

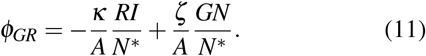

In order to obtain these expressions we assumed that *N*≫*N*\*, which is a well satisfied assumption in most applications. In the following, since *A* is fixed, we will replace the quantities *κ/A* and *ζ/A* by *κ* and *ζ* so that *κ*^−1^ and *ζ*^−1^ can be associated with the typical timescales of diffusion during expansion and shrinking, respectively. In more sophisticated descriptions the influence of the uncertainty of these parameters can be investigated^23^, or its values can be changed drastically to mimic mobility restrictions^5^.

In the present description all terms are expressed as dimensional quantities. In order to simplify the analysis, we introduce dimensionless variables, where all compartments are normalized by *N*. For instance we note 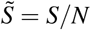, and introduce the same normalization for *B, I, G, R*. This naturally implies *Ñ*\* = *N*\**/N*. Omitting in the following the tildes, since we will only use normalized quantities, we have

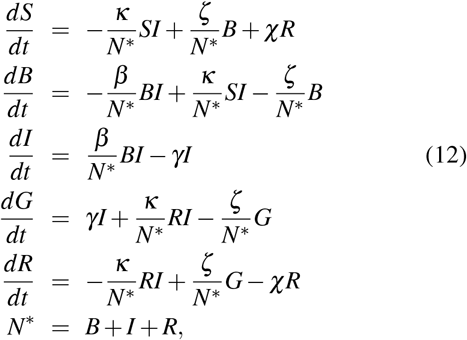

where the division by *N*\*, a quantity which varies in time as the epidemic evolves, is a very important feature of the SBIGR model. By identifying the ratios 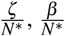 and 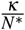 to normalized coefficients of the differential system, this introduces without any ad-hoc or external forcing a time dependence of the otherwise constant coefficients *ζ, β* and *κ*.

Before numerically integrating the model, we first interpret the physical meaning of the 5 model-parameters.

The value of *γ*^−1^ represents the typical duration of the contagious period of an individual and *β* will determine the contagiousness, i.e., the rate at which an infected individual contaminates susceptible individuals. These two quantities determine to a large extent the initial, exponential phase of an epidemic. In particular, their ratio *β/γ* determines the reproduction number in the beginning of an epidemic. This number *β/γ* = *ℛ*_0_(0) ≈ 2, at least in the beginning of the COVID-19 pandemic^24^. The new quantities in the SBIGR model are the growing and shrinking time-scales 1*/κ* and 1*/ζ*, as well as 1*/χ*, if decrease of immunity is taken into account.

The *κ* parameter, associated with the expansion of the blob sets the height of the first epidemic peak. Indeed, the SBIGR dynamics allow to reproduce the saturation of an outbreak before the total population has attained herd-immunity. It is possible to estimate the peak-value of the number of infected individuals analytically (see appendix). The peak is given by

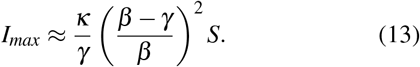

The parameter *ζ* does not appear in this expression, which governs the short-time behavior of the epidemic wave, but is key in determining the long-time dynamics. Indeed, the blob deflates after local herd immunity has been attained, and we show in the appendix that at long times the blob evolves as

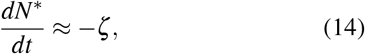

which allows to show that for given *β* and *γ* the typical decay time scales like *T* ∼ *κ/*(*ζγ*).

These ideas are further assessed by numerical integration of the model. We use the PyGom library developped by Public Health England^25^, that makes use of the integrators provided by the SciPy package. We solve the Initial Value Problem with a time-step of one day. Thereto we need the definition of initial conditions and values for the control parameters. For the initial conditions we consider the case of the very beginning of an epidemic where *B*(0) = *G*(0) = *R*(0) = 0, *I*(0) ≡ *I*_0_ = 10^−5^ ≪ 1, and *S*(0) = 1 − *I*_0_. The parameters *β* and *γ* are determined such that *β/γ* = 2. We take *γ* = 0.18 which gives an order of magnitude of the infectious period of approximately 1 week (72% of the individuals have been cured or removed from the *I* compartment after 7 days). These values are of the order of magnitude of the first epidemic wave of the COVID-19 epidemic. We set for the moment *χ* = 0 (persistent immunity).

We illustrate the model with *κ* = 0.002 and *ζ* = 0.00015 in Fig. 3. The most salient feature of this stackplot is the cyclic nature of the epidemic: even in the absence of decay of immunity (*χ* = 0), the SBIGR model, representing the epidemic as an inflating and deflating blob, exhibits well-defined waves, characteristic for pandemics such as COVID-19.

**FIG. 3.**
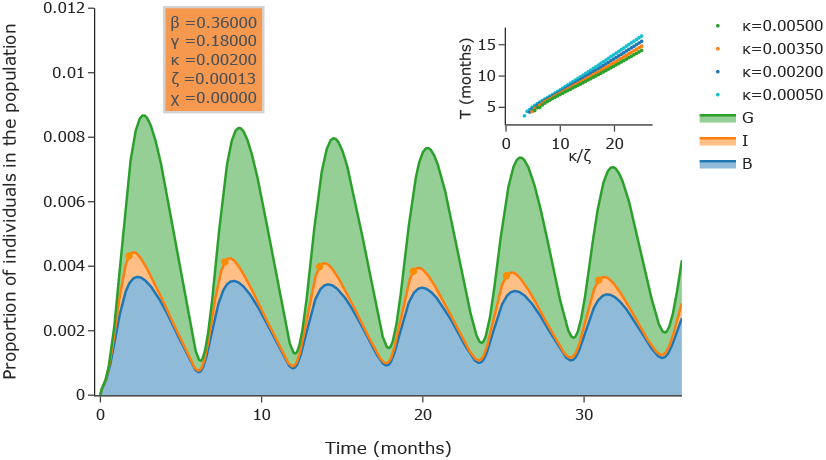
(a) Numerical integration of the SBIGR model [Eqs. (12)]. The figure shows a stackplot of the quantities *B, I, G*. Slowly decaying oscillations are observed for this set of parameters. In the inset it is illustrated that the time-interval *T* between two peaks of the number of infected individuals (indicated by dots in the main plot) is set by the parameter *κ/ζ*.

In order to understand the influence of the parameters on the period of the waves, we have systematically varied *ζ* for different values of *κ* (keeping *κ > ζ*). It is shown in the inset of Fig. 3, as demonstrated in the supplementary materials, that the effective period between successive local maxima of the number of infected is proportional to *κ/ζ*.

A standing question is obviously whether this very simple model shows more than qualitative agreement with a realistic pandemic. However, quantitative comparison over long time-periods is not straightforward. Clearly, in the current pandemic, different countries have used different control strategies such as strict lockdowns. These would to some extent influence the parameters *β* and *κ* of the model, and possibly *ζ*, so that these parameters cannot be chosen strictly constant anymore. This will possibly influence the periodic character of the pandemic, and more certainly modify the relative heights of successive peaks.

Varying the model parameters as a function of time is a common way to reproduce a posteriori the evolution of an epidemic^26^. This would however severely complicate the understanding of the model. It is remarkable that the present model with constant coefficients reproduces saturation of the first wave followed by a close to periodic slowly decaying dynamics. We have investigated different countries and have observed that some countries show a more periodic behavior than others. One of the countries where the data of the number of daily new infected cases is most periodic is South-Africa. We keep *β* = 0.36, *γ* = 0.18 and the linear relation in the inset of Fig. 3 allows to determine the value for *κ/ζ*. Subsequently results for different values of *κ* are shown in Fig. 4.

**FIG. 4.**
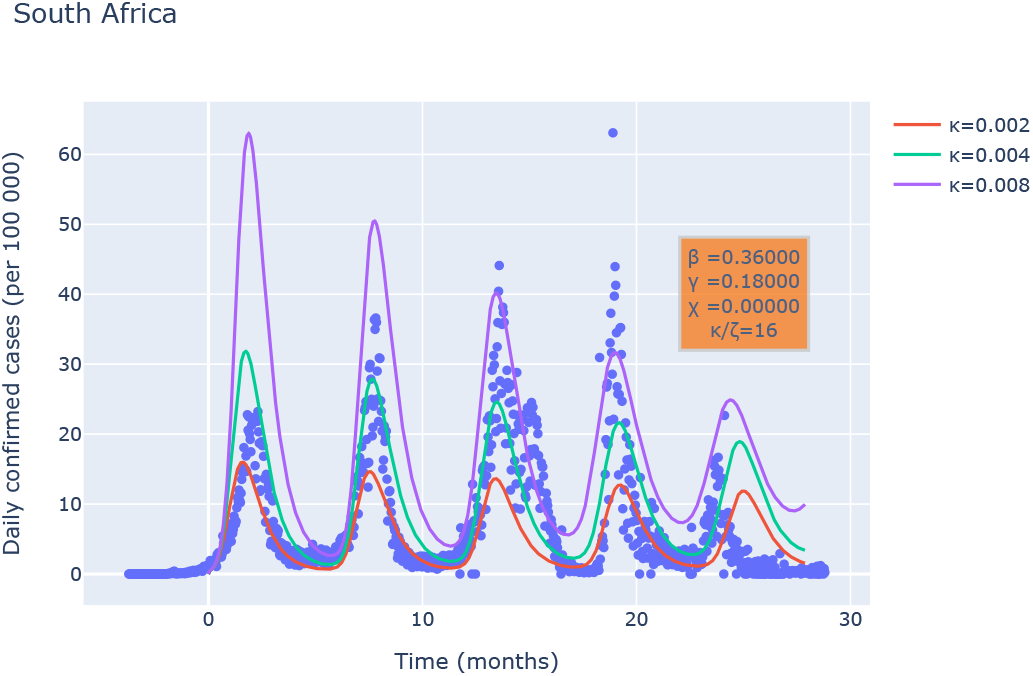
(a) Comparison of the results of the SBIGR model [Eqs. (12)] with data of newly reported cases for South Africa. Results for different values of *κ* are shown for a fixed value of *κ/ζ*.

The most important observation is that the SBIGR model allows to reproduce, for these model-parameters, the correct period of the waves and a good estimate of the order of magnitude of the number of infected individuals. Indeed the exact number is most probably underestimated significantly in the beginning of the epidemic since tests were less available. Furthermore, what our model does obviously not take into account in its simplest form, is the genetic evolution of the virus leading to variants which have different properties, thereby affecting the values of *β, γ*. Nevertheless, even without taking these effects into account, the results are strikingly similar, and we hope that more sophisticated models based on this framework, beyond the scope of the present work, will significantly improve our understanding of the epidemics.

### b. The existence of a limit-cycle

The epidemic waves in Fig. 4 are very pronounced. Their amplitude decays and clearly the number of infected individuals will eventually tend to zero and stay there. Indeed, since the population-size *N* is fixed and lasting immunity is obtained, the epidemic will eventually die out. If we model, as in Fig. 1 the transfer from the *R* to the *S* compartment by a linear transfer term ±*χR*, the natural decay of immunity can be taken into account. This can correspond both to genetic evolution of the virus, or to the evolution of the immune-system of the individuals in the *R*-compartment. We have added, for the same parameters as used to model the evolution of the COVID-19 pandemic in South-Africa, this transfer term and we have varied the value of *χ*. More complicated models could introduce a nonlinearity in this term, or time-variations, mimicking the different immune decline of variants of a virus.

We illustrate, using a phase-space plot in BIG space in Fig. 5, that the dynamics of our model tends to a limit-cycle for *χ > χ*_crit_ = 7.15 × 10^−4^, corresponding to a value 1*/χ*_crit_ ∼ *t*_1*/*2_ ≈ 10^3^ days, the time at which the immunity has decayed for an isolated person to 50%. This value of *t*_1*/*2_ is an order of magnitude larger than estimated for the COVID-19 disease, where *t*_1*/*2_ = *𝒪* (10^2^) days^27^. Considering that in the current pandemic the natural immunity acquired after infection decays with a typical timescale approximately 3 months, we can conclude that, within the scope of our model, the COVID-19 virus will evolve on such a limit-cycle.

**FIG. 5.**
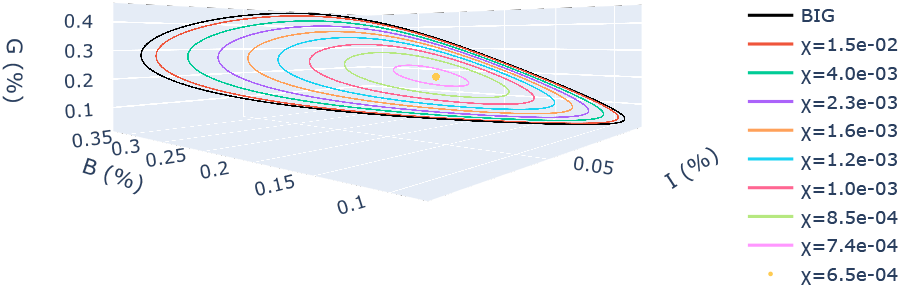
Phase-space plot of the SBIGR model [Eqs. (12)] with decrease of immunity, *χ >* 0. For the current parameters, for values of *χ > χ*_crit_ = 7.15 *×* 10^−4^, corresponding to a typical immunity decrease time *t*_1*/*2_ ≈ 1000 *days* leads to a limit cycle, representing a non-vanishing periodic epidemic. For *χ <* 7.15 × 10^−4^, the system reaches a stable point.

The origin of the limit-cycle can be illustrated analytically by carrying an analysis of the simplified system where we set the quantities *S* and *R* to 1 and 0, respectively. This allows to reduce the model to three ODEs which mimic the beginning of the epidemic described by the SBIGR model. For this reduced system it is straightforward to determine expressions for the fixed-points of the system, and the eigenvalues associated with the linearized system. In the appendix we show that this analysis yields, for fixed *γ* and *β*, values for *κ* and *ζ* which lead to a limit-cycle. As we illustrated in Fig. 5, this limit-cycle is in the full model (with evolving *S, R*) damped for *χ < χ*_crit_.

## Discussion & Conclusion

The main conclusion that can be drawn from the current work is that epidemic waves can be caused by the spatial nature of the spreading of the disease which will, at long times, often be slower than the local saturation. This saturation allows to decrease the local concentration of infected so that the spread slows down. However, this saturation does not, by any means indicate the end of an epidemic, since the deflation of the blob, representing the ensemble of clusters around infected individuals will eventually lead to a situation where a new spread of the disease is possible. The resulting epidemic waves can therefore not be eradicated by a lock-down which is shorter than at least several times *ζ*^−1^. Obviously, the model needs to be refined before precise quantitative predictions can be formulated. Furthermore, even though compartmental models are powerful tools in the study of epidemic spreading, uncertainty in coefficients leads to a large unpredictability at long times^23^. In our opinion the main contribution of such models is therefore the understanding of phenomena and their ability to probe the influence of certain measures. The power of our approach is that, to reproduce the main features of the COVID-19 epidemic (saturation and cyclic dynamics), the SBIGR model does not need to model the effect of social-distancing, finite incubation time, demographic evolution, lock-down, seasonal fluctuations, vaccination, evolution of the virus etc. The complexity of the model remains therefore limited to 5 quantities, evolving according to 5 ODEs with constant parameters, opening a way to efficiently construct more complex models to assess the influence of different factors on epidemic waves.

## Data Availability

All data produced in the present study are available upon reasonable request to the authors

## Data Availability Statements

The data that support the findings of this study are openly available from the Johns Hop-kins University data-base^2^.

## ACKNOWLEDGMENTS

The authors thank Benjamin Pillot (UMR Espace-Dev 228) for developing the Pycovid-19 package^28^ used to access data of the JHU-database. We also thank Simon Bos Sanz for suggesting to add the linear model for the decay of immunity.

## APPENDIX

### Short time behavior: height of the first peak

In order to better understand the influence of the different parameters, we analytically estimate the height of the peak as a function of *κ* assuming *ζ* = 0. The resulting equation for *B* becomes then,

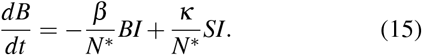

A maximum for *B* is obtained by setting the time-derivative to zero leading to the solution for the maximum of *B* at time *τ*,

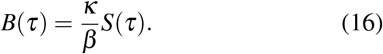

In order to estimate the height of the first peak, we assume exponential increase of both *I* and *G* in this phase. The exponential of *I* is determined by the exponent *β* − *γ* and *G* will increase with the same exponential time-dependence. Indeed, during this phase, we have

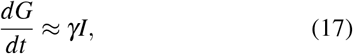

so that, if *I* ∼ exp(*β* − *γ*) ∼ *G*, we have

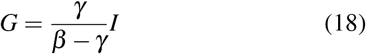

and we assume that this approximation holds until the number of infected individuals peaks. We now set the time-derivative in the equation for *I* equal zero to obtain at this peak,

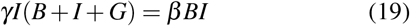

since *I >* 0, and using Eqs. (18) and (16), we have at *t* = *τ*,

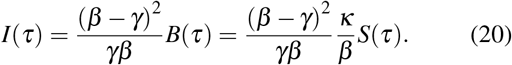

If, finally, we assume that the number of infected peaks approximately at the same time as *B*, and that *S*(*τ*) ≈ *S*(0) = 1, we obtain that the first peak of the number of infected is given by

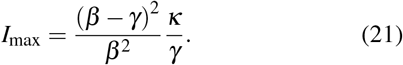

This expression shows thus that the height of the first peak of the epidemic is directly proportional to the value of *κ*.

### Time-scale of the epidemic waves

An estimate of the period of the waves can be obtained by considering the dynamics of the blob. The blob is defined as *N*\* = *B* + *I* + *G*. Summing the equations for *B, I, G* yields

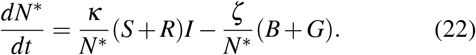

We can assess the long-time evolution of an epidemic wave, when *I* ↓ 0. In this limit, the second term becomes dominant, and *N*^*^ ≈ *B* + *G*. We obtain then the expression,

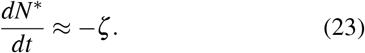

The decay of the blob is therefore linear, to that we can estimate the order of magnitude of the time *T* from the peak of a wave to the beginning of the next wave by

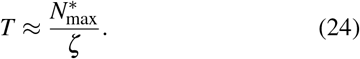

In order to estimate the maximum blob size, we combine the expressions for the maximum of *I, G* and *B* (relations (18), (16) and (20)) to find the expression,

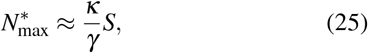

so that the typical decay-time can be estimated, to be

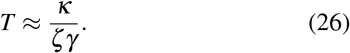

This expression is thus an order of magnitude estimate of the typical decay-time of an epidemic wave, and determines, in the periodic regime, the time-interval between two successive epidemic waves.

### Analysis of the limit-cycle

In order to pinpoint the origin of the oscillatory behavior, we set *S* = 1 and *R* = 0. This yields the simplified system,

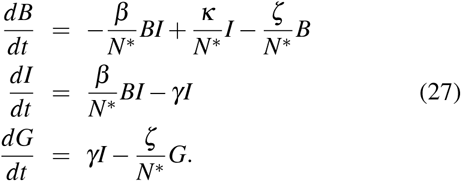

The fixed point of this system is

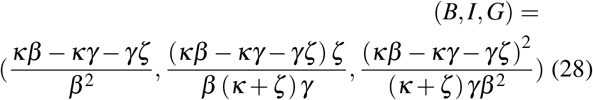

The Jacobian of the system shows that for the parameters used in this study [*β* = 0.36, *γ* = 0.18, *ζ* = 1.25 × 10^−4^, *κ* = 2 × 10^−3^] the fixed point has two complex conjugate eigen values and one real eigenvalue:

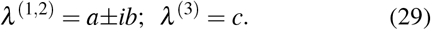

We find in day^−1^ units *a* = 1.32 × 10^−3^, *b* = 0.0357, and *c* = −0.0718. This is representative of an unstable point (*a >* 0), the trajectory around the point focusing on a plane orthogonal to the third eigenvector (*c <* 0) and rotating with a period 2*π/b* = 5.8 months, as observed in Fig. 3.

## Notes

### Competing Interest Statement

The authors have declared no competing interest.

### Funding Statement

This study did not receive any funding

### Summary of Updates

Third revision under consideration for Phys Rev E, notation changed on demand of one referee.

